# A Blood-Based Synaptic Biomarker in Neuronally-Enriched Extracellular Vesicles Predicts Disability Progression in Progressive Multiple Sclerosis

**DOI:** 10.64898/2026.10.07.26364982

**Authors:** Larissa Jank, Madathiparambil KS Kumar, Deepika Joshi, Veronica Minney, Rahel Eskinder, Rohit Thapa, John G. Heller, Angeliki G. Filippatou, Fatemeh Siavoshi, Matthew D. Smith, Christopher Harp, Ibraheem Abioye, Elias S. Sotirchos, Blake E. Dewey, Shiv Saidha, Kathryn C. Fitzgerald, Peter A. Calabresi, Ellen M. Mowry, Pavan Bhargava

## Abstract

Progressive multiple sclerosis (PMS) lacks blood-based biomarkers of the neurodegenerative processes driving disability. We measured synaptopodin, a postsynaptic protein, in neuronal-enriched extracellular vesicles (NEVs) across two independent cohorts: the SPRINT-MS trial (n=184) and a real-world validation cohort (n=268). A one SD increase in baseline synaptopodin was associated with ∼32% lower risk of confirmed disability progression in PMS (p=0.011), with the strongest effect in secondary PMS (SPMS) (∼57% lower odds, p=0.019). This SPMS association was replicated in the validation cohort (∼60% lower odds, p=0.009). Higher baseline synaptopodin was associated with future slower whole-brain atrophy and preserved microstructural integrity across MRI metrics (p=0.007-0.013). Longitudinal changes in NEV synaptopodin were associated with disability worsening (p=0.007) and paralleled MRI microstructural changes. NEV synaptopodin, but not NfL, was associated with confirmed disability progression in PMS participants with both biomarkers measured (HR 0.71 vs 1.12, p=0.007 vs 0.22 for NEV synaptopodin and NfL, respectively) and SPMS (HR 0.58 vs 1.10, p=0.003 vs 0.50). Adding synaptopodin, but not NfL, to a base model improved discrimination for disability progression in SPMS in both cohorts (e.g. validation AUC 0.755 to 0.838, p=0.005). These findings identify NEV synaptopodin as a prognostic and longitudinal, blood biomarker of synaptic integrity in PMS, particularly SPMS, complementing NfL.

## Introduction

Multiple sclerosis (MS) is the leading cause of progressive non-traumatic disability in young adults in developed countries^1^. Although disease-modifying therapies modulating peripheral immune cell functions have significantly improved outcomes in relapsing forms of MS (RMS), treatment options for progressive MS (PMS) remain limited. In contrast to RMS, PMS pathology is mainly driven by CNS intrinsic processes, including neurodegeneration, CNS-restricted inflammation, and demyelination with failed remyelination.^2^ While these processes occur throughout the CNS, gray matter (GM) pathology is a major driver of cognitive decline and long-term disability in PMS.^3,4^ A key barrier to developing treatments for PMS has been the lack of biomarkers that reflect progressive pathology.

Serum neurofilament light chain (NfL) is the leading blood biomarker in MS. NfL is a non-specific marker of neuroaxonal injury and closely tracks inflammatory disease activity, with strong evidence, particularly in RMS cohorts.^5–8^ However, in PMS (and arguably also in RMS^9^), circulating NfL levels increase in response to both acute focal injury and slower, chronic neurodegeneration. The small changes occurring with disease progression can fall below a measurable threshold or can be masked by other axonal damage^6^. Therefore, the utility of NfL alone as a biomarker of neurodegeneration in PMS is limited. There is a strong need for blood biomarkers of gray matter pathology, a key driver of disability progression, which, when combined with markers such as NfL, would enable a more complete assessment of the underlying processes driving disability progression.

MS has increasingly been described as a synaptopathy, in which reactive microglia aberrantly prune synapses and contribute to accelerated neurodegeneration.^10,11^ Synaptic dysfunction and synaptic loss represent candidate disease processes for biomarkers in MS. In the CSF, synaptic proteins such as SNAP-25 and neurogranin decline with longer disease duration and are associated with brain atrophy.^12^ Furthermore, recent studies using SV2A positron emission tomography (PET) have linked reduced synaptic density in cortical gray matter to worse cognitive performance.^13,14^

Extracellular vesicles (EVs) provide a blood-accessible compartment that carries protected protein cargo, and neuronal enrichment increases representation of CNS proteins in plasma-derived EV fractions.^15^ CNS tissue EVs isolated from MS brain and MS animal models show pronounced alterations in synaptic protein cargo, often stronger than changes measured in matched tissue.^16,17^ These findings strongly support neuronal-enriched EVs (NEVs) as a platform for synaptic biomarkers in MS.

Synaptopodin is a postsynaptic, spine-associated protein and a marker of synaptic structure.^18^ Previous studies from our group have linked NEV synaptopodin to MS status and shown that longitudinal changes in NEV synaptopodin are associated with brain atrophy and retinal thinning.^19,20^ While these studies support a rationale for investigating synaptopodin as an NEV biomarker, they included modest sample sizes and focused primarily on relapsing disease (n=15 PMS participants in both studies).

There is a critical need for biomarkers of MS progression. This study aims to address this gap by advancing an NEV synaptic biomarker. We hypothesized that NEV synaptopodin reflects ongoing synaptic pathology in PMS and would therefore (1) predict future clinical and MRI progression, (2) longitudinally concurrently track disability and MRI progression and respond to neuroprotective treatment, and (3) provide information complementary to NfL.

To test these hypotheses, we measured synaptopodin in NEVs from plasma using a harmonized workflow across two independent cohorts: a discovery cohort from the SPRINT-MS randomized trial^21^ (n=184), and an independent real-world validation cohort enriched for progressive disease (n=268). The SPRINT-MS trial was an informative discovery cohort because it included people with PMS, had standardized MRIs (including DTI and MTR) and regular plasma sampling at baseline, week 48, and week 96, and met its primary endpoint of slowing whole-brain atrophy (though ibudilast did not significantly slow disability progression a secondary endpoint), also allowing testing of treatment responsiveness of the biomarker to ibudilast. The independent real-world validation cohort was selected because it enabled us to test whether the findings from the controlled clinical trial could be generalized to clinical practice, where patient populations are more heterogeneous, MRI is acquired on multiple scanners with varying protocols, and follow-up visits occur at irregular intervals.

## Results

### Cohort and characteristics

The discovery cohort (SPRINT-MS) included 184 participants with PMS, including 104 with PPMS and 80 with SPMS. Compared with PPMS, SPMS participants included a higher proportion of females (72.5% vs 42.3%), a longer disease duration (16.4 ±8.8 vs 7.6 ±7.2 years, p <0.001), and a higher baseline Expanded Disability Status Scale (EDSS) score (5.7 ±0.9 vs 5.1 ±1.1, p <0.001). Age did not differ significantly between PPMS and SPMS participants (55.7 ±6.8 vs 56.9 ±7.1 years, p=0.11). The placebo and ibudilast groups were balanced for age, sex, disease duration, and baseline EDSS scores. Race differed between treatment arms (90.4% white in placebo vs 97.8% in the Ibudilast arm, p=0.035).

The validation cohort included 268 participants with MS, including 90 people with RMS and 178 with PMS. Compared with RMS, PMS participants showed a longer disease duration (15.9 ±9.3 vs 10.4 ±6.9 years, p <0.001) and higher baseline EDSS scores (4.5 ±1.6 vs 2.5 ±1.3, p <0.001). Within the PMS participants in the validation cohort, more people had SPMS than PPMS (78.6% vs 21.4%), people with SPMS had a longer disease duration than people with PPMS (17.5 ±9.0 years vs 10.4 ±8.0, respectively; p <0.001). Disease-modifying therapy utilization differed between RMS and PMS participants, as well as between those with PPMS and SPMS. Table 1 lists the full demographic and clinical characteristics of both cohorts.

**Table 1.:**
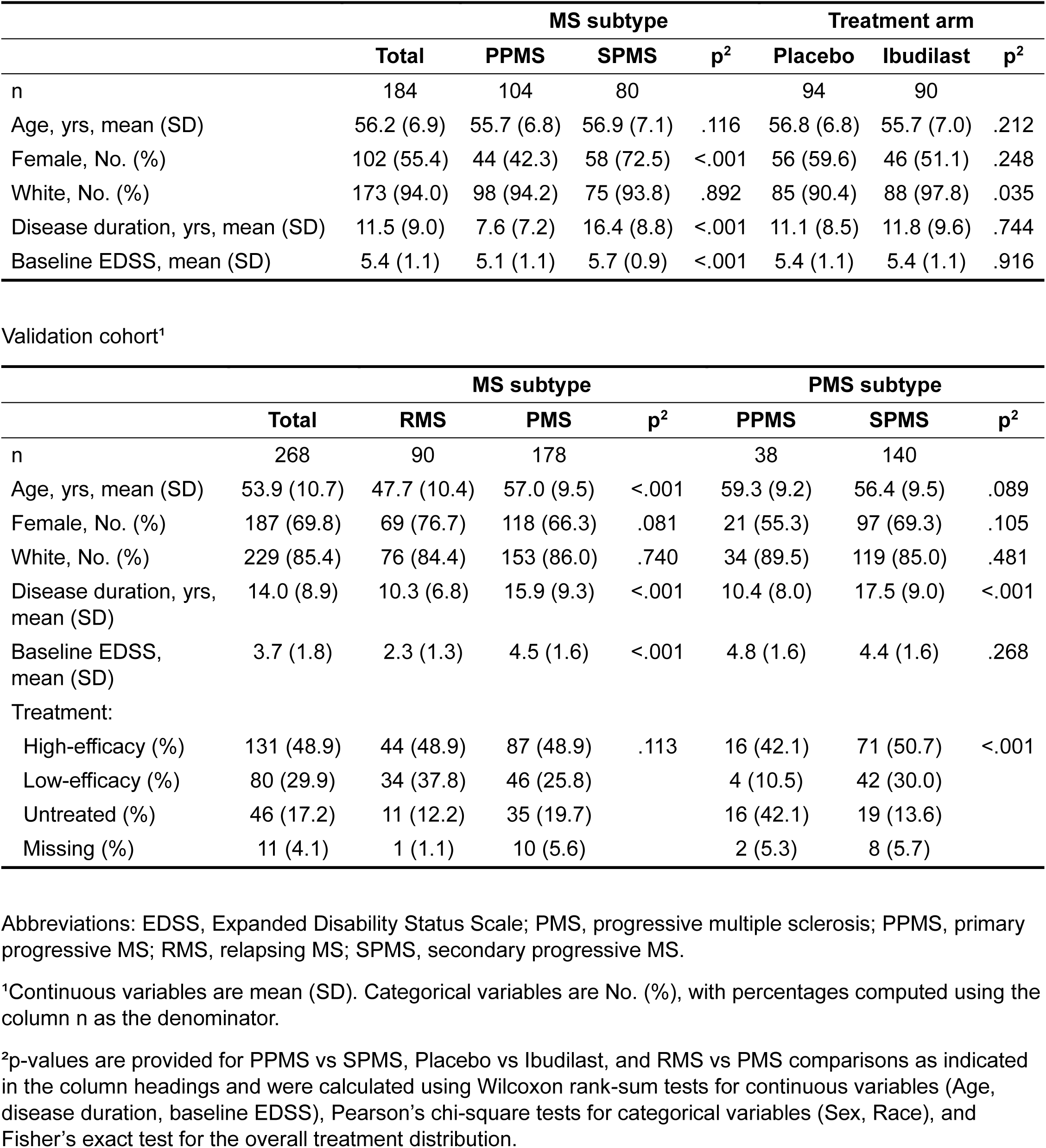
Baseline demographic characteristics in the discovery and validation cohorts.

### Assay characterization

Total EVs were isolated from defibrinated plasma by polymer-based precipitation (ExoQuick), followed by enrichment of NEVs using L1CAM immunoprecipitation (workflow in Fig. 1a). Negative-stain electron microscopy of NEV isolates showed particles in the expected small-EV size range alongside some stained background, consistent with co-isolation of non-vesicular plasma components that can accompany precipitation-based workflows. Nanoparticle tracking analysis (NTA) showed comparable median particle sizes across plasma, total

**Fig. 1.:**
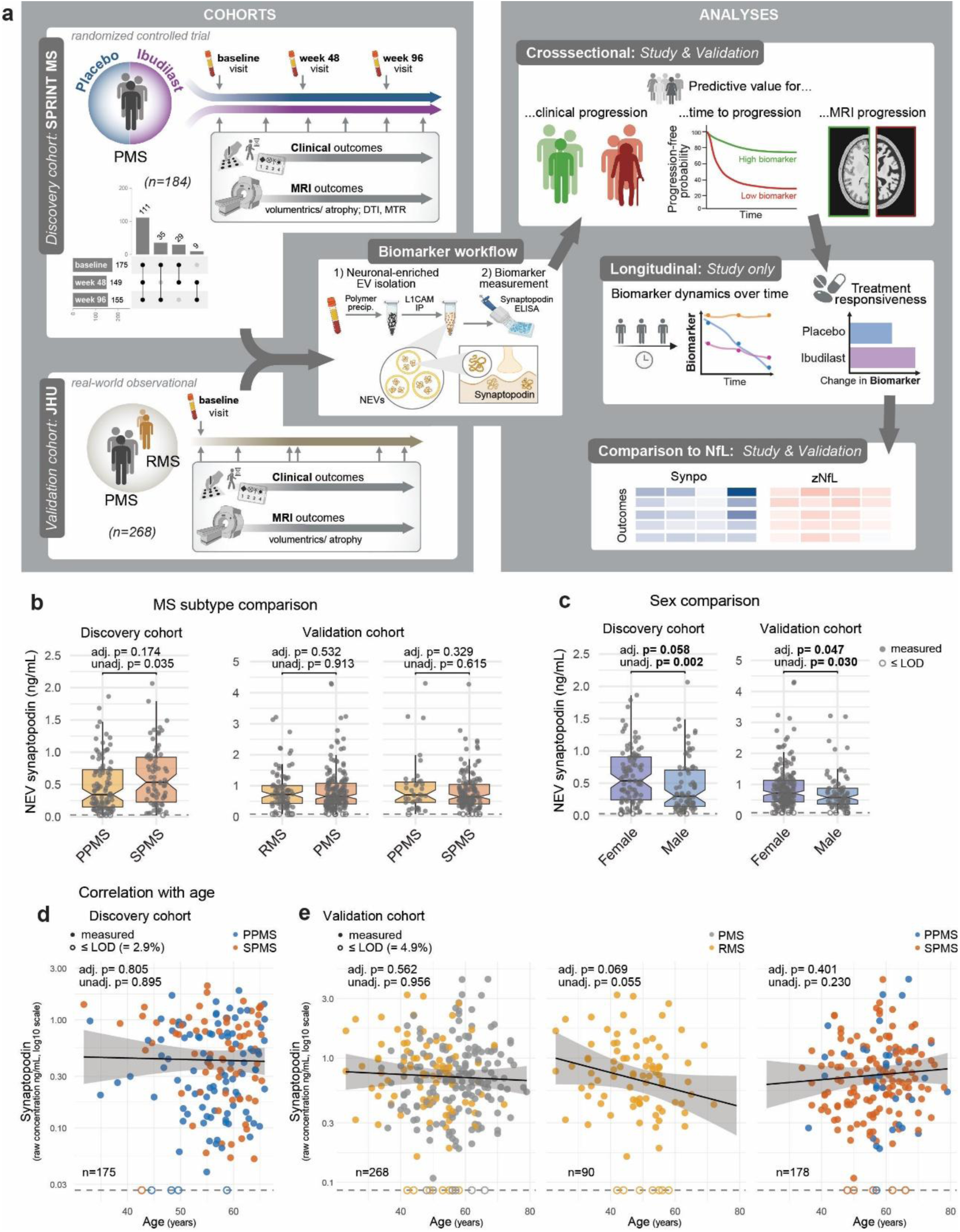
Study design and baseline NEV synaptopodin comparisons in the discovery and validation cohorts. **(a)** Schematic overview of the study. Left: cohort sampling schedules for the discovery and validation cohorts, including an UpSet plot showing the completeness of NEV synaptopodin measurements across time points for the discovery cohort. The validation cohort only includes baseline measurements. Middle: overview of NEV isolation workflow and synaptopodin quantification. Right: analysis overview, including baseline prognostic analyses, longitudinal analyses, and comparison with NfL. Notched boxplots of the baseline NEV synaptopodin concentration by MS subtype **(b)** and sex **(c)** in the two cohorts. Unadjusted comparisons used Wilcoxon rank-sum tests. Adjusted p-values are from ANCOVA of synaptopodin z-scores adjusting for age, sex, race, MS/PMS subtype, and treatment arm (excluding the respective grouping variable). Association of baseline NEV synaptopodin with age in the **(d)** discovery and **(e)** validation cohort are shown with subgroups indicated by color and limit of detection (LOD) indicated by the dashed line and open symbols. The line and band (95% CI) show the covariate-adjusted (same covariates as in panel b) associations between age and synaptopodin z-scores with back-transformed results displayed as concentrations. P-values are for the adjusted age associations and unadjusted Spearman correlations. **Sample size:** Discovery cohort: n=175 with baseline synaptopodin measures; Validation cohort: n=268.

EVs, and L1CAM+ EVs (all ∼100nm), indicating that enrichment primarily altered composition and yield rather than particle size distribution. Protein concentration decreased stepwise from plasma to total EVs and further to NEVs. In contrast, particle concentration was similar between plasma and total EVs but markedly lower in NEVs, consistent with selection of a smaller, antigen-defined particle subfraction. The particle-to-protein ratio was higher in total EVs compared to NEVs, suggesting that immunoprecipitation isolates may be relatively protein-rich. This could be explained by immunoprecipitation reagents (e.g., antibody/BSA) or by free/cleaved L1CAM, which might coprecipitate with the EVs. Synaptopodin concentration was higher in NEVs than in total EVs, consistent with enrichment of neuronal cargo in the L1CAM+ fraction.

In the discovery cohort, NEV synaptopodin was quantified at least once (baseline, week 48, or week 96) for all 184 participants. We retained participants with missing timepoints when the available measurements were sufficient for the respective analyses (data completeness summarized in the UpSet plot, Fig. 1a). In the validation cohort, all 268 participants had baseline NEV synaptopodin measurements. Values at or below the assay limit of detection (LOD) were infrequent across the discovery cohort visits (baseline 2.9%; week 48 3.4%; week 96 3.9%) and in the validation cohort (4.9%).

### NEV synaptopodin levels are higher in females with MS

There were no marked differences in NEV synaptopodin levels across MS subtypes (Fig.1b). In the discovery cohort, SPMS participants had higher baseline NEV synaptopodin than PPMS in unadjusted comparisons, but this difference was attenuated after covariate adjustment. One plausible explanation for this is the pronounced sex imbalance between PMS subtypes in this cohort, with a higher proportion of females in SPMS than PPMS (72.5% vs 42.3%). In the validation cohort, PMS participants did not have a significant difference in proportion of females between SPMS and PPMS.

In both cohorts, females had higher baseline NEV synaptopodin than males (Fig.1c). After adjusting for age, race and MS subtype and (where applicable) treatment, females had 35.6% higher levels than males in the discovery cohort (-1.1%-85.9% CI) and 24.7% higher levels in the validation cohort (0.3%-54.9% CI). Baseline NEV synaptopodin levels were not associated with age in either cohort across subgroups (Fig. 1d-e), with the exception of a statistically non-significant trend toward lower levels with increasing age in people with RMS in the validation cohort (unadjusted p=0.06; covariate-adjusted p=0.07). There were no notable differences in baseline NEV synaptopodin levels by race in either cohort, by ibudilast/placebo treatment status in the discovery cohort (Fig. S1f), or by disease-modifying therapy (DMT) class in the validation cohort (Fig. S1g). Supplementary Table S1 lists NEV synaptopodin levels across demographic groups.

### Baseline NEV synaptopodin predicts disability progression and MRI outcomes in SPMS in the discovery cohort

In participants in the discovery cohort (people with PMS) with baseline NEV synaptopodin and outcome data (n=175), higher baseline synaptopodin was associated with a lower likelihood of confirmed disability progression (OR 0.636 [0.441–0.918], p=0.016; Fig. 2a and supplementary table S2). This effect was predominantly driven by people with SPMS (OR 0.426 [0.208–0.870], p=0.019). In this subgroup, each one standard-deviation increase in baseline NEV synaptopodin was also associated with reduced odds of EDSS progression (OR 0.37 [0.19-0.75], p=0.006), EDSSplus progression (OR 0.54 [0.29-1.01], p=0.053), and timed-25-foot-walk (T25FW) progression (OR 0.37 [0.18-0.72], p=0.004). In analyses using continuous disability trajectories in SPMS (Fig. 2b and supplementary table S3.1) the directionality of the correlations aligned: higher baseline synaptopodin was associated with slower worsening of T25FW and nine-hole peg test (9HPT) trajectories in the SPMS subgroup, although neither of these analyses reached statistical significance (in SPMS association with T25FW ρ=-0.221 [-0.432–0.013], p=0.064 and for 9HPT ρ=-0.197 [-0.407–0.033], p=0.093).

**Fig. 2.:**
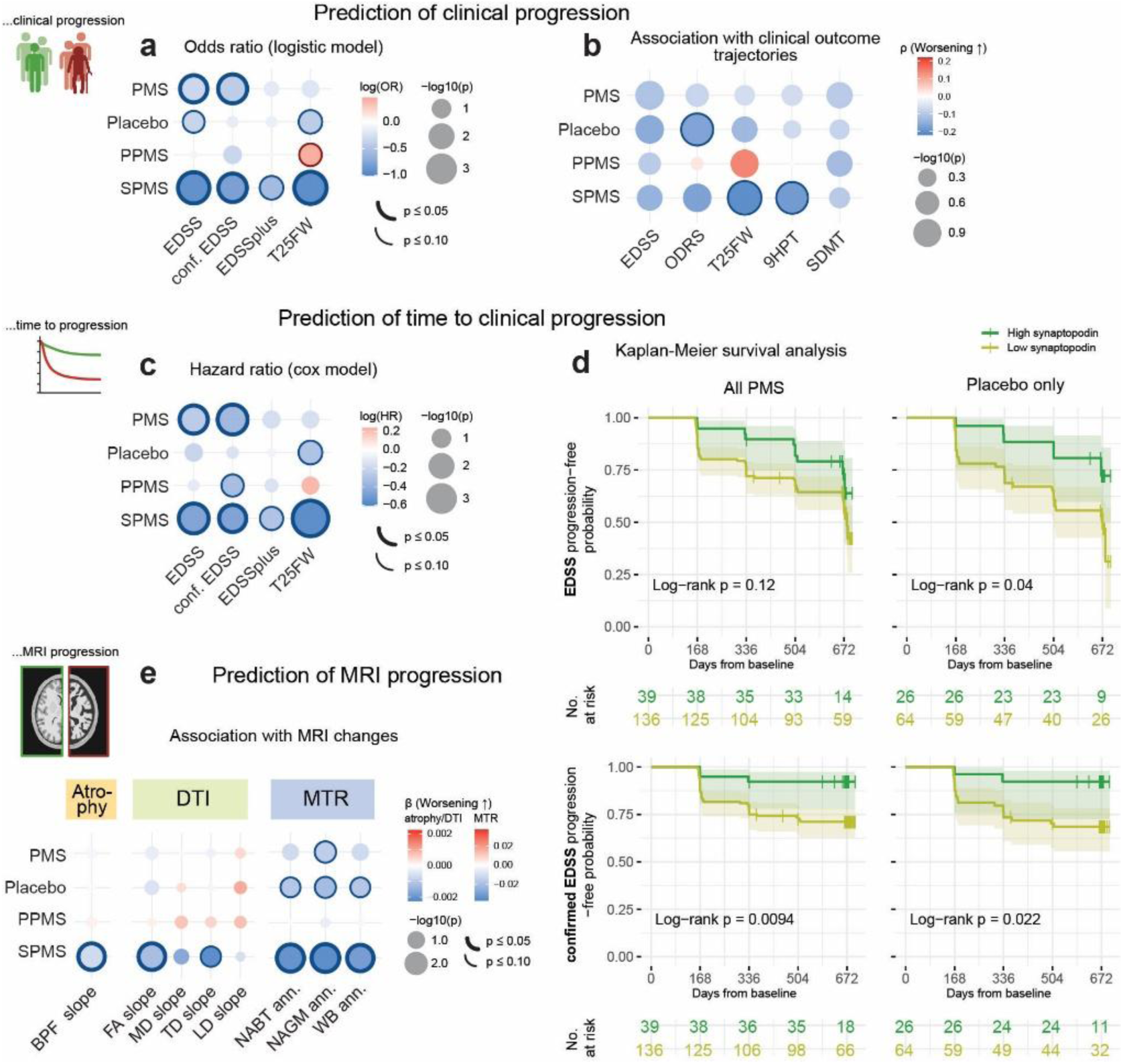
Baseline NEV synaptopodin is associated with clinical progression, worsening trajectories, and MRI change in the discovery cohort. **(a)** Dot-matrix plot of odds ratios (OR) for binary progression outcomes across subsets. Logistic models included age, sex, race, PMS subtype, and treatment arm, along with the corresponding baseline clinical measure for each outcome (placebo models omitted treatment arm). Dot color represents log(OR) (positive/red indicates higher odds of progression), dot size represents −log10(p-value), and outlines indicate P≤0.05 (thick) or 0.05<P≤0.10 (thin). **(b)** Dot-matrix plot of covariate-adjusted partial Spearman correlations (ρ) between baseline NEV synaptopodin and continuous outcome trajectories across subsets. Partial correlations were adjusted for the same covariates as above. Dot color represents ρ (positive/red indicates worse progression); dot size and outlines match panel a. **(c)** Dot-matrix plot of hazard ratios (HR) for time-to-progression outcomes across subsets. Cox models included the same covariates as in panel a. Dot color represents log(HR) (positive/red worse); dot size and outlines match panel a. **(d)** Kaplan-Meier curves for the progression-free probability stratified by baseline NEV synaptopodin (high/dark green vs low/light green) using a ROC-derived Youden cutoff optimized for confirmed EDSS progression and the full cohort (all PMS). Shaded bands represent 95% confidence intervals; number-at-risk tables and log-rank p-values are shown below. **(e)** Dot-matrix plot of covariate-adjusted β coefficients showing the associations between baseline NEV synaptopodin and longitudinal MRI outcomes (Brain parenchyma fraction (BPF)/ Diffusion Tensor Imaging (DTI)/ Magnetization Transfer Ratio (MTR) change measures) across subgroups. Models included age, sex, race, PMS subtype, and treatment arm, as applicable, along with the corresponding baseline MRI measure. Dot color represents β (with positive/red indicating worsening of the MRI measure); dot size and outlines match panel a. **Sample size:** for most analyses n=175/90/101/74 (Full/Placebo/PPMS/SPMS); details in supplementary tables.

In time-to-progression analyses (Fig. 2c and supplementary table S2), higher baseline synaptopodin predicted longer time to confirmed disability progression (HR 0.678 [0.503–0.914], p=0.011). This effect, too, was driven by people with SPMS, in whom the biomarker predicted a 39.3% reduced risk of confirmed EDSS progression (HR 0.61, p=0.032), and a 45.5% reduced risk of T25FW progression (HR 0.55, p<0.001). Kaplan-Meier analyses (Fig. 2d) confirmed that participants with higher baseline synaptopodin had longer disability progression-free survival than those with lower baseline synaptopodin in the full cohort (log-rank p=0.0094) and in a sensitivity analysis limited to placebo-treated participants (p=0.022).

Higher baseline NEV synaptopodin was associated with slower brain atrophy and preserved microstructural integrity (Fig. 2e and supplementary table S3.2). These associations were the strongest in the SPMS subgroup, where baseline synaptopodin was associated with slower whole brain atrophy (change in brain parenchymal fraction [BPF], p=0.010), changes in fractional anisotropy (FA) (p=0.009), and evolution of magnetization transfer ratio (MTR) measures (p=0.013–0.007). In the full PMS cohort, associations were directionally consistent, but most did not reach statistical significance.

### The real-world validation cohort confirmed NEV synaptopodin as a biomarker of clinical progression and brain atrophy in PMS

In participants from the independent real-world validation cohort with baseline synaptopodin and outcome data (n=188-201 in the full cohort and n=123-131 for PMS, depending on the outcome), higher baseline synaptopodin was again associated with lower odds of confirmed disability progression within people with PMS (0.511 [0.288– 0.907], p=0.022) (Fig. 3a, and supplementary table S2). As in the discovery cohort, these effects were predominantly driven by the SPMS subgroup (0.396 [0.198–0.792], p=0.009). A similar pattern was observed for EDSSplus progression (p=0.014), as well as in analyses assessing EDSS scores as a continuous variable (Fig. 3b and supplementary table S3.1). Here, higher baseline synaptopodin was associated with slower EDSS worsening in SPMS (ρ=-0.208 [-0.388–-0.012], p=0.038). A sensitivity analysis also including participants with up to two missing EDSS functional scores (vs. only one missing in the primary analysis), to maximize inclusion, showed consistent results for confirmed EDSS progression in PMS (n=137 vs. 123, OR 0.512 [0.292–0.898], p=0.020) and SPMS (n=110 vs. 100; OR 0.455 [0.241–0.859], p=0.015; HR 0.552 [0.339–0.900], p=0.017) (supplementary table S2).

**Fig. 3.:**
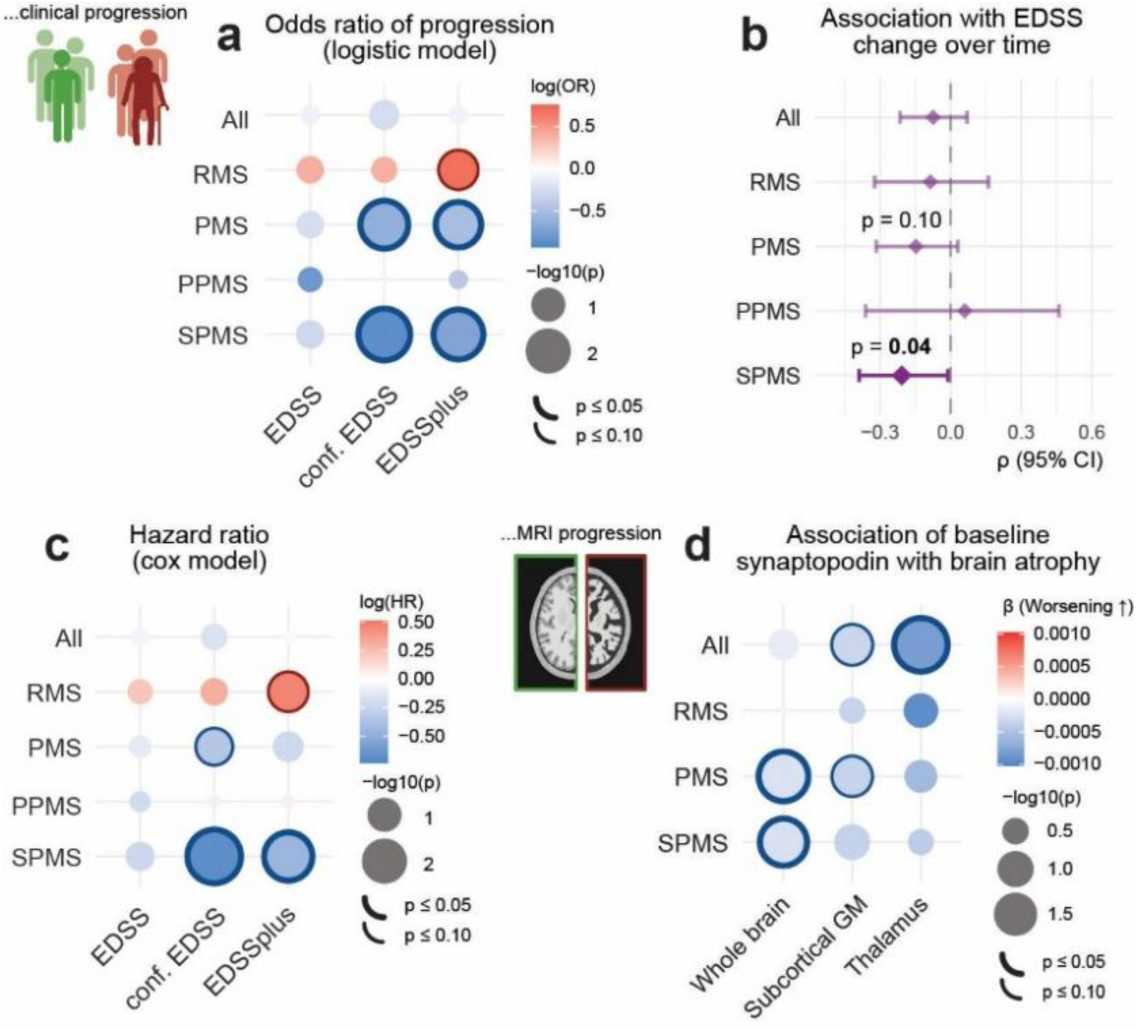
Baseline NEV synaptopodin is also associated with clinical progression, worsening trajectories, and brain atrophy in the validation cohort. **(a)** Dot-matrix plot of odds ratios (OR) for binary progression outcomes across subsets. Logistic regression models included age, sex, race, baseline EDSS, and MS/PMS subtype (as applicable i.e. for All: RMS vs PMS and within PMS: PPMS vs SPMS). Dot color represents log(OR) (positive/red indicates higher odds of progression), dot size represents −log10(p-value), and outlines indicate P≤0.05 (thick) or 0.05<P≤0.10 (thin). **(b)** Forest plot of covariate-adjusted partial Spearman correlations (ρ) between baseline NEV synaptopodin and continuous EDSS trajectories across subsets. Partial correlations were adjusted for the same covariates as in panel a. The x-axis represents ρ (with negative ρ indicating associations with slower EDSS progression). **(c)** Dot-matrix plot of hazard ratios (HR) for time-to-progression outcomes across subsets adjusting for the same covariates as in panel a. Dot color represents log(HR) (positive/red worse) with dot size and outlines matching panel a. **(d)** Dot-matrix plot of covariate-adjusted β coefficients showing the associations between baseline NEV synaptopodin and longitudinal MRI outcomes across subgroups. Models included age, sex, race, MS/PMS subtype (as applicable, see panel a), and the corresponding baseline MRI measure. Dot color represents β (with positive/red indicating associations with faster brain atrophy); dot size and outlines match panel a. **Sample size:** Panels a, b, and c (All/RMS/PMS/PPMS/SPMS): n=188/65/123/23/100. Panel d: MRI slopes n=120/50/71/13/57.

In time-to-progression analyses (Fig. 3c, and supplementary table S2), higher baseline NEV synaptopodin predicted a longer time to confirmed disability progression in SPMS (HR 0.479 [0.285–0.805]; p=0.006). Besides confirmed EDSS progression, NEV synaptopodin also predicted EDSSplus progression in SPMS (p=0.020). Kaplan-Meier stratification using the discovery cohort Youden cutoff did not statistically significantly separate progression curves in the validation cohort (supplementary Fig. S2a), however an exploratory post-hoc cohort-specific optimized cutoff yielded a significant separation in SPMS (supplementary Fig. S2b).

The associations between baseline NEV synaptopodin and MRI brain atrophy measures in the real-world validation cohort were similar to those in the discovery cohort, despite the MRIs being based on post-acquisition harmonized clinical scans from different instruments (Fig. 3d and supplementary table 3.2). NEV synaptopodin was associated with slower whole-brain atrophy in both the overall PMS group (p=0.030) and people with SPMS (p=0.047). The associations with subcortical and thalamic atrophy were directionally aligned but did not reach statistical significance in these subgroups. In the full cohort, however, the association with thalamic atrophy was significant (p=0.014), possibly reflecting the greater statistical power of the entire cohort.

### Changes in NEV synaptopodin track clinical and MRI progression in the discovery cohort

The SPRINT-MS trial included longitudinal plasma draws at weeks 48 and 96 after baseline, enabling exploratory longitudinal analysis of the biomarker. Participant-level annualized synaptopodin slopes were estimated from all available measurements for participants with measurements at ≥2 timepoints. Slopes were estimable for 184 participants, of whom 9 did not have baseline measurements and were thus excluded from those analyses, adjusted for baseline biomarker levels. The slopes did not significantly differ by sex, race, or PMS subtype and were not correlated with age (Supplementary Fig. S3a-b and supplementary table S4).

To test if changes in synaptopodin reflect changes in clinical disease progression, we assessed synaptopodin slopes in progressors and non-progressors defined according to clinical measures (Fig. 4a). In the full cohort (n=175), people with MS (PwMS) without disability progression, defined by stable or improving multidomain overall disability response score (ODRS), had significantly more increasing synaptopodin trajectories (ANCOVA β=-0.0346 [-0.0596–-0.0095], p=0.007). A similar trend was observed when progression was defined by EDSS alone, though this did not reach statistical significance (ANCOVA β=-0.0217 (-0.0436–0.0002), p=0.052). Progression defined by 20% worsening of T25FW or 9HPT was directionally consistent but not statistically significant, while no association was observed for cognitive worsening defined by symbol digit modalities test (SDMT; supplementary Fig. S3 and supplementary table S5). When analyzing ODRS as a continuous variable (Fig. 4b and supplementary table S6), increasing NEV synaptopodin over time was associated with slower ODRS worsening (ρ=-0.153 [-0.295–-0.005], p=0.043), with a stronger association observed in just the placebo group (ρ=-0.215 (-0.404–-0.008), p=0.042).

**Fig. 4.:**
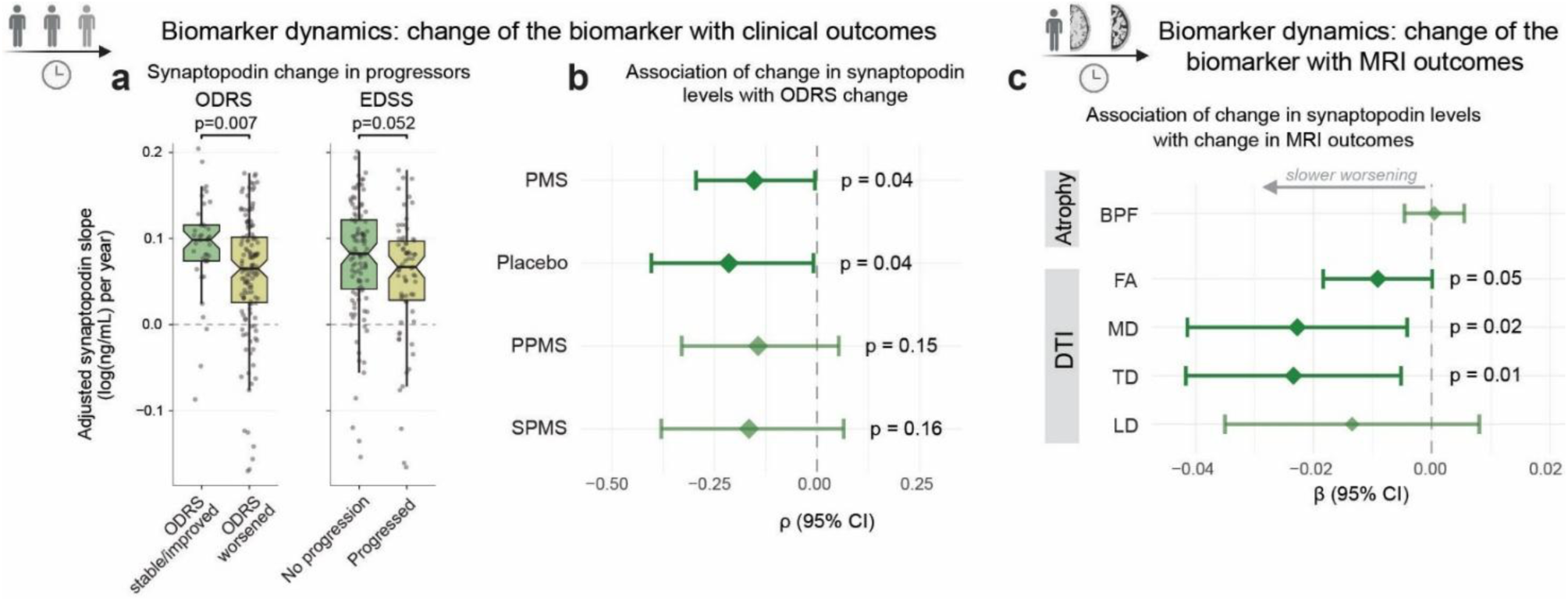
Longitudinal NEV synaptopodin change reflects disability progression and MRI change. **(a)** Boxplots of the adjusted NEV synaptopodin change over time by ODRS (stable/improved vs. worsened; left panel) and EDSS progression (progressed vs. not progressed; right panel) status in PMS participants (adjusted for age, sex, race, PMS subtype, treatment arm, baseline NEV synaptopodin z-score, and baseline EDSS). Adjusted p-values were derived from ANCOVA. **(b)** Forest plot of covariate-adjusted partial Spearman correlations (ρ) between NEV synaptopodin change over time and ODRS scores across subsets. Partial correlations were adjusted for the same covariates as in panel a. The x-axis represents ρ (with negative ρ indicating associations with less ODRS worsening). **(c)** Forest plot of covariate-adjusted partial Spearman correlations (ρ) between NEV synaptopodin change over time and MRI measures in PMS. Partial correlations were adjusted for age, sex, race, PMS subtype, treatment arm, baseline NEV synaptopodin z-score, and the corresponding baseline MRI measure. The x-axis represents ρ (with negative ρ indicating associations with slower MRI measure worsening). **Sample size:** For all Panels (Full/Placebo/PPMS/SPMS): n=175/90/101/74, except MTR MRI measures here n=153 for the full cohort.

Increase in NEV synaptopodin was also associated with slower MRI progression (Fig. 4c and supplementary table S6). Across the full cohort (n=175), greater increase in synaptopodin was associated with slower diffusion MRI worsening (mean diffusivity slope p=0.017; transverse diffusivity p=0.012, and fractional anisotropy p=0.053). However, NEV synaptopodin change was not associated with changes in BPF or in MTR.

Exploratory analyses testing if NEV synaptopodin trajectories differed by ibudilast treatment arm did not show a statistically significant effect of ibudilast on longitudinal synaptopodin both in linear mixed-effects models (Supplementary Fig. S3d and Supplementary Table S7) and when comparing precalculated annualized patient-level NEV synaptopodin slopes by ANCOVA (Supplementary Fig. S3e).

Collectively, these analyses show that longitudinal changes in NEV synaptopodin capture ongoing accumulation of disability and microstructural injury on MRI.

### NEV synaptopodin captures a disease axis distinct from NfL in PMS

Analyses to compare the performance of NEV synaptopodin to the more established biomarker NfL as a biomarker of disease progression, were restricted to PMS participants with both NEV synaptopodin and NfL z-scores available (discovery cohort: n=171; validation cohort: n=147). In both the discovery and validation cohorts, synaptopodin and NfL z-scores were not significantly correlated with one another in PMS overall or in the SPMS subgroup (discovery r=−0.02, validation r=0.10; Fig. 5b).

**Fig. 5.:**
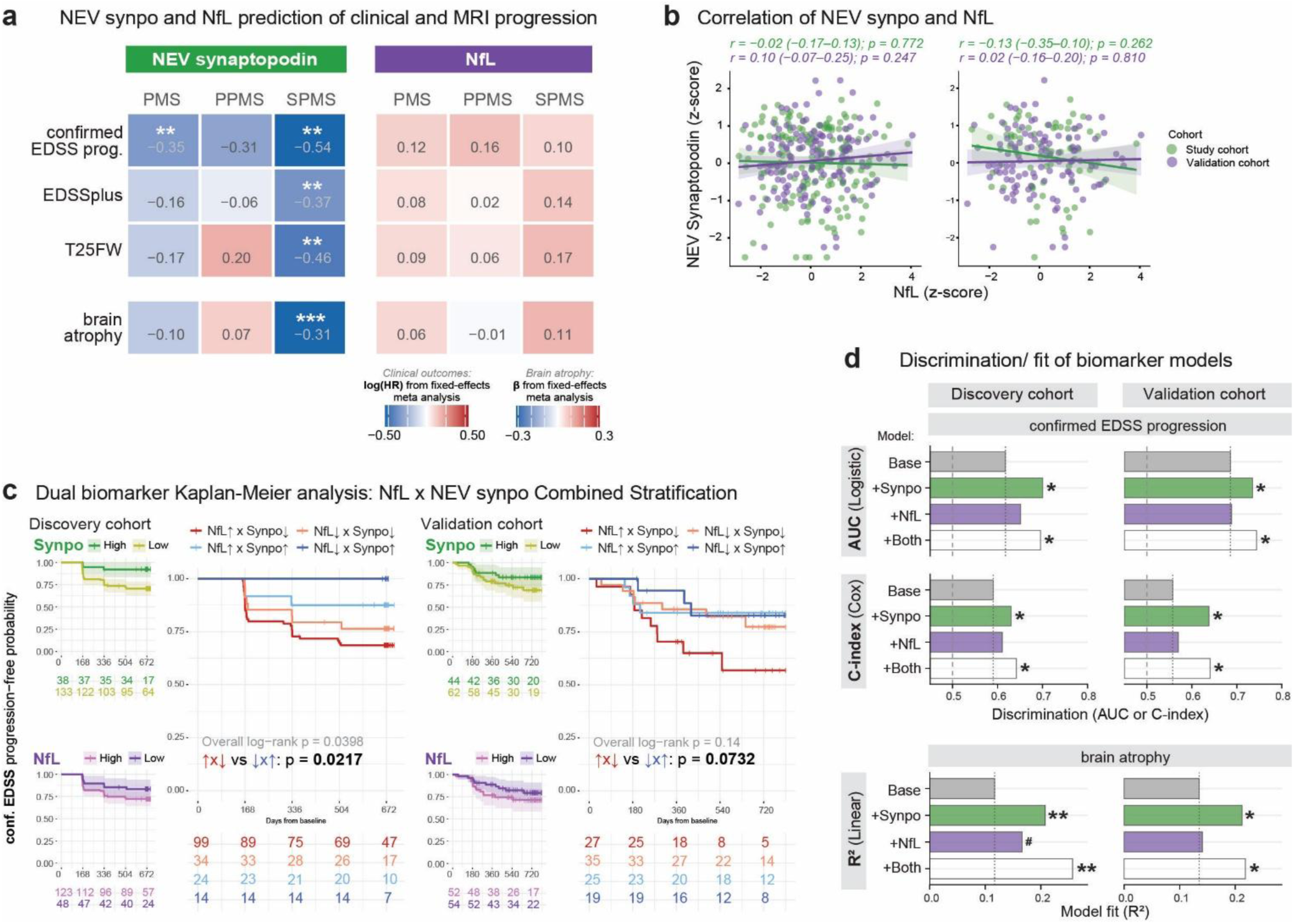
Comparison of NEV synaptopodin and NfL as biomarkers of disease progression. **(a)** Heatmap of pooled fixed-effects meta-analysis for NEV synaptopodin and NfL across clinical progression outcomes and brain atrophy in PMS, PPMS, and SPMS subgroups. Tile color and effect sizes represent pooled log(HR) from Cox models for clinical outcomes and pooled standardized linear β for whole-brain atrophy, where red indicates worse progression or faster atrophy. Models were adjusted for age, sex, race, PMS subtype (as applicable), and baseline clinical outcome or baseline MRI measures; discovery cohort models were additionally adjusted for treatment arm. Significance is annotated with: #P≤0.10, *P≤0.05, **P≤0.01, and ***P≤0.001. **(b)** Scatterplots of the correlation between baseline NEV synaptopodin and NfL in PMS (left) and SPMS (right) and cohort-specific regression lines with 95% CI bands with Pearson correlation coefficients (r) with 95% CIs, p-values, and sample sizes shown above the plots. **(c)** Kaplan-Meier curves for disability progression-free probability using single-and dual-biomarker stratification based on ROC-derived Youden cutoffs for NEV synaptopodin and NfL.Smaller panels show individual biomarker curves for NEV synaptopodin (high/dark green vs. low/light green) and NfL (high/light purple vs. low/dark purple) separately; the larger panel shows the four-group dual-biomarker stratification (NfL↑Synpo↓/NfL↑Synpo↑/NfL↓Synpo↓/NfL↓Synpo↑); Left: Discovery cohort; Right: Validation cohort (PMS subgroup). Overall log-rank p-values and pairwise comparisons between the most discordant groups (NfL↑Synpo↓ vs. NfL↓Synpo↑) are shown. Number-at-risk tables are shown below each plot. **(d)** Horizontal bar charts showing discrimination performance of base, base+NEV synaptopodin, base+NfL, and base+NEV synaptopodin+NfL models for confirmed disability progression (AUC from logistic models and C-index from Cox models) and whole-brain atrophy (R² from linear models; base and combined bars approximated from partial R²), shown separately for the discovery and validation cohorts. Bar colors represent the base model (gray), base+synaptopodin (green), base+NfL (purple), and base+synaptopodin+NfL (white). Dashed lines mark the chance discrimination, and the dotted lines show the base model value. The significance of each biomarker addition relative to the base model was assessed using the likelihood ratio test (#P≤0.10, *P≤0.05, **P≤0.01, ***P≤0.001). The base model included age, sex, race, and baseline EDSS. Analyses were restricted to SPMS participants with both NEV synaptopodin and NfL available. Sample sizes: Discovery cohort (PMS/PPMS/SPMS) clinical and brain atrophy n=171/98/73; Validation cohort clinical n=106/21/85 and brain atrophy n=65/13/52.

Baseline synaptopodin, but not NfL, significantly predicted subsequent confirmed disability progression in the meta-analysis (Fig. 5a and supplementary table S8; synaptopodin: OR=0.61 [0.45–0.84], p=0.002; HR=0.71 [0.55–0.91], p=0.007; NfL: OR=1.14 [0.91–1.42], p=0.247; HR=1.12 [0.93–1.35], p=0.218). These results were consistent on a cohort level. NfL was not significantly associated with confirmed EDSS progression in either cohort, while synaptopodin predicted confirmed EDSS progression in the discovery cohort (n=171 with both NfL and synaptopodin measurements and clinical outcome data; OR p=0.015, HR p=0.010), with directionally aligned, albeit not statistically significant, results in the validation cohort (n=106; OR p=0.065, HR p=0.326), likely reflecting reduced power.

The most pronounced and consistent findings were in the SPMS subgroup, where synaptopodin was associated with confirmed EDSS progression (OR=0.43 [0.26–0.73], p=0.001; HR=0.58 [0.41–0.84], p=0.003), EDSSplus composite progression (OR=0.50 [0.32–0.79], p=0.003; HR=0.69 [0.53–0.91], p=0.008) and T25FW in the meta-analysis, though the association with T25FW was mainly driven by the discovery cohort. NfL was non-significant for all these outcomes in SPMS (all p>0.16).

Lower baseline synaptopodin was associated with faster whole-brain atrophy in SPMS in the meta-analysis (β=−0.00038 [-0.00064−-0.00013], p=0.003) and in both cohorts individually (discovery p=0.009; validation p=0.043). A significant association was also present in the meta-analysis of PMS overall (β=−0.00021 [−0.00041 to −0.00001], p=0.038), driven by the validation cohort. Baseline NfL did not predict whole-brain atrophy in SPMS, PPMS, or PMS overall (p>0.073).

When participants were stratified into four groups based on baseline synaptopodin and NfL levels (combining the Youden cutoffs identified earlier for synaptopodin and new Youden cutoffs for NfL), the discordant groups showed the most extreme progression-free survival in both the discovery and validation cohorts (Fig. 5c). PwMS with high synaptopodin and low NfL progressed slowest while those with low synaptopodin and high NfL progressed fastest. These two groups were significantly different from each other in the discovery cohort and showed a consistent trend in the validation cohort (high NEV synatopodin/ low NfL vs low NEV synaptopodin/ high NfL: discovery cohort p=0.040, validation cohort p=0.140 for PMS overall).

To investigate whether combining the biomarkers would improve their predictive performance, we tested whether adding the biomarkers separately and in combination to a base model (age, sex, race, baseline EDSS) improved the AUC and C-Index of models predicting confirmed disease progression and brain atrophy in SPMS (Fig. 5d). Adding baseline synaptopodin significantly improved prediction of confirmed disability progression in both cohorts (AUC: discovery 0.618 to 0.701, p=0.014; validation 0.755 to 0.838, p=0.005; C-index: discovery 0.590 to 0.630, p=0.046; validation 0.550 to 0.662, p=0.018). Adding synaptopodin also significantly improved prediction of whole-brain atrophy in SPMS in both cohorts (marginal R²: discovery 0.117 to 0.208, p=0.007; validation: 0.135 to 0.212, p=0.035). Adding NfL alone did not significantly improve discrimination for either outcome in either cohort (all p>0.09, except for a trending improvement in the whole-brain atrophy model, p=0.059). Adding both biomarkers did not meaningfully improve discrimination beyond synaptopodin alone.

These results suggest that the two biomarkers have distinct advantages in progressive disease: synaptopodin is a robust predictor of disability progression, particularly SPMS, while combining both biomarkers identifies prognostic subgroups not captured by either marker alone.

## Discussion

This study provides evidence of a synapse-linked and EV-derived blood biomarker for disability prediction in PMS, thereby addressing the critical need for biomarkers of MS progression. Across two independent cohorts, higher baseline NEV synaptopodin predicted reduced likelihood of subsequent disability worsening and MRI progression in people with PMS, especially those with SPMS. Longitudinal changes in NEV synaptopodin tracked clinical and MRI progression in the SPRINT-MS trial. In direct comparison, synaptopodin demonstrated advantages over NfL in predicting disability and MRI outcomes. However, stratifying PwMS by both biomarkers identified subgroups at particularly high or low risk of progression, an effect not captured when combining them into a single statistical model, suggesting the two markers contribute non-redundant prognostic information. Taken together, these findings establish NEV synaptopodin as a blood-accessible marker of progressive disease biology that complements rather than duplicates existing biomarkers.

The rationale for measuring synaptopodin, a postsynaptic protein required for spine apparatus formation and synaptic plasticity^18^, in NEVs was based on evidence from EV tissue studies, CSF biomarkers, and synaptic density imaging. In MS brain tissue and experimental autoimmune encephalomyelitis spinal cord, our prior work demonstrated that CNS tissue-derived EVs show pronounced downregulation of synaptic proteins, with changes often more pronounced in EVs than in matched tissue homogenates^16,17^. In CSF, declining levels of synaptic proteins have been linked to longer disease duration and reduced brain volumes^12^. SV2A PET imaging has directly associated lower cortical synaptic density with worse cognitive performance in MS.^13,14^ EVs offer a practical advantage for translating these observations into blood: protein cargo is protected within vesicles, and cell-type-specific proteins present at low concentrations in plasma can be enriched by surface-marker immunocapture.^15^ Building on this biological rationale, we examined whether baseline NEV synaptopodin levels could serve as a biomarker for MS disability progression.

The prognostic associations between baseline NEV synaptopodin and subsequent disability worsening replicated across the two independent cohorts despite fundamental differences in their design, demographics, and treatment exposure. Consistent findings across logistic, time-to-event, and continuous slope analyses strengthen confidence in the association. The effect sizes were very similar across cohorts, with each SD increase in baseline synaptopodin associated with a ∼32% lower risk of disability progression in people with PMS in both cohorts (32.2% and 31.1%, discovery and validation respectively). Confirmed disability progression was significant for both OR and HR in the discovery cohort but only OR in the validation cohort, most likely reflecting reduced power for confirmation analyses in the smaller validation cohort PMS group (n=123 vs n=175). Supporting this, EDSSplus, which captures worsening across multiple functional domains, was significant for both OR and HR in the real-world setting. This pattern, therefore, probably reflects differences in outcome sensitivity across study designs rather than inconsistency in the underlying signal. A sensitivity analysis restricted to placebo-treated participants in the discovery cohort showed consistent effect directions across all analyses, with several reaching statistical significance despite reduced power, suggesting that the results were not driven by treatment effects. Some clinical endpoints did not reach statistical significance, likely due to low event rates and the relatively short follow-up time to monitor disease progression (∼2 years for both cohort). Significant associations with continuous disability trajectories in both cohorts suggest a meaningful prognostic signal that is likely to strengthen with longer follow-up.

Interestingly, in both the discovery and the validation cohorts, the association was mostly driven by the SPMS subgroup, where effect sizes were larger than in PMS overall and confirmed disability progression reached independent statistical significance with almost identical effect sizes (∼57% and ∼60% lower odds, in the discovery and validation cohort respectively). One possible explanation is statistical: the PPMS subgroup was smaller, especially in the validation cohort, and the analysis may have been underpowered to detect comparable effects. An alternative explanation is that the synaptic pathology captured by NEV synaptopodin is more pronounced in PwMS with longer accumulated disease burden and prior inflammatory activity (during the relapsing stage), which in our cohorts was the SPMS group. Based on the current data, we cannot distinguish between these explanations. However, if the subtype pattern reflects a real difference in biomarker associations across PMS subgroups, this would have implications for patient selection in trials targeting synaptic protection.

Longitudinal changes in synaptopodin paralleled clinical disease course with participants who remained stable showing increases in synaptopodin compared to those that progressed. This was supported by continuous analyses in which a greater increase in NEV synaptopodin over time was associated with slower ODRS worsening in PMS overall and most strongly in placebo-treated participants. However, the short follow-up with only three timepoints limits our ability to determine whether synaptopodin changes precede clinical worsening. Future studies with more frequent sampling over longer periods are needed to establish the temporal relationship between changes in synaptopodin and disease progression.

Higher baseline synaptopodin predicted slower future brain atrophy and microstructural injury, supporting and extending the clinical findings. The two cohorts differed in their MRI acquisition: the discovery cohort used standardized protocols with fixed acquisition parameters that included DTI and MTR sequences, while the validation cohort used mostly clinically acquired multi-site images with varying protocols and parameters that required harmonization. In the discovery cohort, baseline synaptopodin predicted slower whole-brain atrophy in SPMS. These associations were confirmed in the validation cohort, despite the greater measurement noise inherent to harmonized real-world MRI data, with slower whole-brain atrophy in SPMS and PMS overall, slower thalamic atrophy in the whole cohort, and a non-significant trend for subcortical gray matter atrophy in PMS. Brain atrophy and synaptic pathology are both recognized features of neurodegeneration in progressive MS, and thalamic atrophy specifically has been linked to the loss of synaptic proteins in CSF in MS^12^.

Microstructural findings in the discovery cohort further support the associations between atrophy and baseline synaptopodin. DTI fractional anisotropy (FA) and MTR associations were predominantly observed in SPMS, mirroring the clinical results. FA has been shown to correlate with clinical disability in MS and reflects subtle changes in tissue microstructure, such as loss of axonal and myelin organization, that precede or occur independently of macroscopic volume loss^22,23^. The association between baseline synaptopodin and FA in SPMS therefore suggests that synaptopodin captures tissue damage at a more subtle microstructural level, beyond the macroscopic volume loss reflected by atrophy measures. For MTR, significant associations were observed across all three measures in SPMS within the discovery cohort, and consistent trends in placebo-treated participants. MTR primarily reflects myelin integrity^24^ rather than synaptic pathology directly. Therefore, the association between synaptopodin and MTR measures suggests that synaptopodin might capture broader tissue damage in progressive disease extending beyond synaptic pathology alone. In the longitudinal synaptopodin analyses (discovery cohort only), changes in NEV synaptopodin were selectively associated with DTI metrics but not with MTR measures. This pattern may reflect the different tissue properties captured by these techniques: DTI metrics better reflect tract integrity and axonal damage, which might be more closely linked to synaptic pathology than the more myelin-sensitive MTR.

Beyond its prognostic utility, direct comparison with NfL helps define what synaptopodin uniquely captures as a biomarker. NEV synaptopodin and NfL were not significantly correlated in either cohort, suggesting they capture independent biological information. The difference in the prognostic performance of the two biomarkers is consistent with the biology of each marker: NfL is established as a marker of acute axonal injury and inflammatory disease activity in RMS and is responsive to disease-modifying therapies.^5–8,25^ In PMS NfL provides less prognostic information about chronic disability accumulation as a standalone marker.^6,26^ Synaptopodin, by contrast, appears to capture neurodegenerative processes that drive disability accumulation, which likely explains its superior performance for disability outcomes in SPMS. This was further supported by the discrimination analyses, in which adding synaptopodin to a clinical base model significantly improved the prediction of confirmed disability progression and brain atrophy in SPMS in both cohorts independently, with AUCs reaching 0.838 in the validation cohort. Adding NfL alone did not improve model discrimination for any of these outcomes in either cohort. Comprehensive biomarker assessment in PMS may therefore require a combination of several biomarkers, for example, a combination of synaptopodin tracking neurodegenerative progression with synaptic pathology, NfL monitoring inflammatory activity and axonal damage, and markers such as glial fibrillary acidic protein (GFAP) capturing CNS-compartmentalized inflammation.

NEV synaptopodin levels were consistently higher in females than in males across both cohorts, aligning with findings in animal studies showing stronger hippocampal synaptopodin expression in female mice.^27^ Interestingly, in PMS, men experience faster disability accumulation and more extensive gray matter atrophy despite lower inflammatory activity.^28,29^ Therefore, higher synaptopodin levels in females might also reflect a greater synaptic reserve or neuroprotective mechanisms^30^. Regardless of biological interpretation, the consistency of this sex effect indicates that clinical application of NEV synaptopodin might require sex-stratified reference ranges or sex-adjusted interpretation thresholds.

Certain limitations affect the interpretation of this study. Samples at or below the limit of detection occurred in 2.9-4.9% of participants, and mean synaptopodin concentrations differed across cohorts, likely explaining why the Kaplan-Meier cutoff derived from the discovery cohort did not significantly separate progression curves in the validation cohort. These limitations highlight the need for assay standardization and cross-cohort normalization strategies before clinical use. The L1CAM enrichment strategy, while effective at increasing the synaptopodin signal, remains controversial.^15,31,32^ Besides L1CAM expression outside the CNS, free L1CAM in plasma is of concern since it may coprecipitate with EVs. In our study, the particle-to-protein ratio in NEV isolates was lower than in total EVs, suggesting protein-rich co-isolates that may include non-vesicular components. Clinical translation will require: standardization of preanalytical parameters^33^ such as plasma collection and storage protocols, automation of isolation protocols (for less hands-on time and higher throughput), evaluation of alternative neuronal enrichment markers, and implementation of more sensitive assays if yield-reducing protocol changes need to be made. Beyond technical improvements, longitudinal studies with extended clinical, MRI, and biomarker follow-up (5-10 years) are needed to determine whether synaptopodin changes precede clinical worsening and to capture progression events that take a longer time to manifest. Further, mechanistic validation by correlating changes in plasma NEV synaptopodin with postmortem synaptic pathology or SV2A PET would strengthen the biological interpretation. Lastly, our tissue EV proteomics identified numerous dysregulated synaptic proteins. Testing some of the most strongly dysregulated candidates in the future may yield even more significant associations.

In conclusion, this study establishes NEV synaptopodin as a blood-based biomarker that predicts and tracks disability progression in PMS. By capturing disability progression and tissue integrity and outperforming NfL for these outcomes, NEV synaptopodin fills a critical gap in the PMS biomarker field. While technical refinements and validation in cohorts with even longer follow-up times are needed, these findings demonstrate the potential of NEV synaptopodin to improve patient stratification and disease monitoring in clinical trials and potentially in clinical practice.

## Methods

### Study Population

The discovery cohort consisted of participants enrolled in the Phase 2 randomized, double-blind, placebo-controlled SPRINT-MS trial (NCT01982942)^21^, which evaluated Ibudilast versus placebo in progressive MS. Besides the trial inclusion criteria, inclusion in our study was further restricted to people with MS (PwMS) who had sufficient plasma samples available for NEV synaptopodin quantification. Of 184 included participants (PPMS n=104, SPMS n=80; placebo n=94, Ibudilast n=90), baseline measurements were available for 175 individuals, with longitudinal measurements at week 48 (n=149) and week 96 (n=155); 111 participants had measurements at all three time points, 64 had two (including baseline), and 9 had two follow-up measurements without a baseline. The trial enrolled participants at 28 sites across the United States within the NIH NeuroNEXT network between November 2013 and June 2014. Participant protection was provided through a central Institutional Review Board at Massachusetts General Hospital. All participants provided written informed consent before any study procedures. The trial was conducted in accordance with the Declaration of Helsinki.

The validation cohort included PwMS from a real-world observational cohort recruited at the Johns Hopkins Multiple Sclerosis Precision Medicine Center of Excellence. Eligible participants were aged 18-75 years with a diagnosis of MS by the 2017 McDonald criteria, able to attend visits every 6 months for 3-5 years, able to undergo 3T MRI, and enrolled in the Johns Hopkins Precision Medicine Center biobanking initiative. Participants with an additional neuroinflammatory or neurodegenerative disorder, a comorbidity precluding safe sample handling, or active cancer or cytotoxic chemotherapy within the preceding 2 years were excluded. Inclusion in the present analysis further required suitable banked plasma for NEV synaptopodin quantification. Plasma was collected between January 2021 and February 2023, with clinical follow-up through August 2024. The cohort consisted of 268 participants with baseline synaptopodin measurements: 90 with relapsing MS (RMS), 38 with PPMS, and 140 with SPMS. Only baseline biomarker measurements were available for this cohort. All participants in the cohort provided written consent in accordance with the Declaration of Helsinki, and the study was approved by the Johns Hopkins Institutional Review Board (IRB00237681).

### NEV Isolation and characterization

NEVs were isolated as described previously^34^. Briefly, plasma was defibrinized by incubation with thrombin, followed by polymer-based precipitation (System Biosciences, #EXOQ20A-1) and immunoprecipitation using biotinylated anti-L1CAM antibodies (clone: 5G3, Thermo Fisher Scientific, #13-1719-28) and streptavidin-coated beads (Thermo Fisher Scientific, #53116). Protein concentration was determined by bicinchoninic acid assay according to the manufacturer’s protocol (Fisher Scientific, #PI23227). Particle size distribution and concentration were characterized by nanoparticle tracking analysis on a ZetaView Nanoparticle Tracker (Particle Metrix GmBH) with the ZetaView software (8.05.14 SP7) as described previously^16^. Briefly, after calibration of the instrument with 100nm diameter beads (Thermo Scientific, #4310A), filtered PBS-diluted samples were injected and recorded at 488nm (Acquisition settings: Temperature: 23°C; camera sensitivity: 80; Shutter speed: 100; Focus: autofocus; frame rate: 30/sec; Analysis parameters: minimum brightness 20, maximum particle size 1000nM, minimum particle size 10nM). Transmission electron microscopy was carried out as described previously^17^. In brief, NEVs were adsorbed onto glow-discharged carbon-coated copper grids (Electron Microscopy Sciences, #CF400-CU-UL). Grids were washed and negatively stained with 1% uranyl acetate. Vesicular morphology and size range were confirmed by imaging at 80 kV on a Hitachi HT7600 transmission electron microscope.

### Synaptopodin Quantification

Synaptopodin concentrations in NEV samples were quantified by ELISA (LSBio, #LS-F21202-1) performed in duplicate according to the manufacturer’s instructions, with optical density read on a FLUOstar Omega microplate reader (BMG LABTECH). Within each plate, a four-parameter logistic curve was fitted to convert optical density readings to concentrations. Duplicate optical densities were averaged prior to curve fitting. A plate-specific background was subtracted from all readings. Samples below the limit of detection were imputed as half the limit of detection. All sample processing and quantification was performed blinded to clinical outcomes, treatment status, and MS subtype in both cohorts, from sample receipt through NEV isolation, ELISA quantification, and standard curve fitting. Samples were identified only by anonymized IDs.

### Clinical Outcome Definitions

In the discovery cohort clinical assessments were performed every 24 weeks over approximately two years. Relapse visits were excluded from all progression analyses. Disability progression was defined as follows: an increase from baseline of ≥1.5 points if baseline EDSS was 0, ≥1.0 points if baseline was 0.5–5.5, or ≥0.5 points if baseline was >5.5. Confirmed disability progression required confirmation at a visit ≥20 weeks after the initial progression date. Since the discovery cohort had EDSS visits every 24 weeks, a confirmation at ≥20 weeks equated to confirmation at the next scheduled visit, allowing for visit-window variability (same definition as in the original trial^21^). T25FW and 9HPT progression was defined as a ≥20% worsening from baseline. SDMT progression was defined as a decrease of ≥8 points from baseline.^35^ Low-contrast visual acuity (LCVA, 2.5% binocular) decline was defined as a ≥7-letter decrease. For the composite EDSSplus^36^ endpoint, composite progression across EDSS, T25FW, or 9HPT (dominant and non-dominant) was defined as the minimum time-to-event across domains. The overall disability response score (ODRS)^37^ was computed as the difference between the number of worsening and improving indicators across EDSS, T25FW, 9HPT (dominant and non-dominant), SDMT, and LCVA. Per-subject continuous outcome slopes were estimated using linear mixed-effects models with random intercepts and slopes.

In the validation cohort, the clinical baseline assessment was defined as the clinical visit closest to the biomarker collection date, within ±90 days. The participants were followed prospectively as part of routine clinical care, with additional regular EDSS assessments approximately every 6 months. The median between-visit interval was 27.4 weeks (IQR 25.5–32.4 weeks), roughly comparable to the discovery cohort 24 week visit schedule. All observations up to 2.5 years were included (median follow-up time 1.99 years [IQR 1.51–2.17 years]). Assessments in the same month as a recorded relapse were excluded, with subsequent months excluded based on documented relapse duration. EDSS scores with 2 or more missing functional scores were excluded (except for the sensitivity analysis in supplementary table S2, which excluded participants missing 3 or more functional scores). The same progression thresholds, composite endpoint definitions, and continuous slope models were applied as in the discovery cohort.

### MRI Acquisition and Processing

In the discovery cohort, MRIs were acquired at weeks 0, 24, 48, 72, and 96. The MRI measures were obtained from the SPRINT-MS imaging analyses as previously described^38–40^. Patient-specific longitudinal slopes for brain parenchymal fraction and diffusion tensor imaging metrics (fractional anisotropy, mean, transverse, and longitudinal diffusivity; averaged across hemispheres) were estimated using unadjusted linear mixed-effects models (random intercepts and slopes for time in years, ≥2 time points required). Magnetization transfer ratio (MTR) in normal-appearing brain tissue, normal-appearing gray matter, and whole brain was summarized as the annualized change from baseline to week 96.

In the validation cohort, a subset of participants had structural MRIs available from research and clinical visits across multiple scanners (only 3D scans were included). Scans were processed through a standardized pipeline that applied deep learning-based super-resolution, longitudinal registration with cross-timepoint harmonization, lesion filling, and atlas-based brain substructure segmentation as described previously^41^. Volumes were normalized by each patient’s median total intracranial volume across visits. Outcomes were annualized percentage change rates for whole-brain atrophy, subcortical gray matter, and thalamic volume. The baseline scan was selected as the scan closest to the biomarker collection date (30 days before and up to 6 months after baseline biomarker measurement). A minimum post-baseline follow-up of 6 months was required (median follow-up time 1.98 years [IQR 1.52–2.18 years]), and participants whose scan time was predominantly accrued before the biomarker collection date were excluded.

### Statistical Analyses

Synaptopodin concentrations (ng/mL) were natural log-transformed and z-scores were determined for each participant within cohorts. Per-subject longitudinal slopes were estimated for the discovery cohort using measurements from at least two or more time points using linear mixed-effects models with random intercepts and random slopes for time. Unadjusted group comparisons of synaptopodin z-scores or per-subject slopes were performed using Wilcoxon rank-sum or Kruskal-Wallis tests. Adjusted comparisons used ANCOVA, with age, sex, race, MS subtype, and treatment arm (discovery cohort) as covariates. For slope models, baseline synaptopodin z-score was added as an additional covariate.

Associations between baseline NEV synaptopodin z-scores and binary clinical progression endpoints were assessed using logistic regression and Cox proportional hazards models, adjusted for age, sex, race, baseline outcome, MS subtype, and treatment arm (discovery cohort). Follow-up time was additionally included in logistic models. Associations with continuous clinical trajectories were assessed using partial Spearman rank correlations, with the same covariates as for the logistic and Cox models. Associations of the biomarker with MRI atrophy rates were assessed by linear regression with the same covariates. All models were fitted separately in prespecified subgroups (PMS, PPMS, SPMS and Placebo only in the discovery cohort and All MS, RMS, PMS, PPMS and SPMS for the validation cohort).

For Kaplan-Meier analyses, participants were dichotomized into high and low NEV synaptopodin z-score groups using an optimal cutoff determined by maximizing the Youden index from a ROC analysis of the progression outcome. For the discovery cohort (which only included PMS participants), the cutoff was derived from the full cohort, and for the validation cohort (which included both PMS and RMS participants) it was derived from the PMS subgroup. Differences between survival curves were assessed using the log-rank test.

The effect of ibudilast on longitudinal NEV synaptopodin trajectories was assessed using a linear mixed-effects model with NEV synaptopodin z-scores as the outcome. The treatment effect was modelled as a treatment-by-time interaction, with random intercepts per participant. Models were adjusted for age, sex, race, and MS subtype. Subgroup analyses in PPMS and SPMS used the same model structure, but without MS subtype as a covariate. For these analyses, all participants with at least one post-baseline measurement were included.

Analyses comparing synaptopodin and NfL were restricted to PMS participants with both biomarkers available (discovery cohort n=171; validation cohort n=147). NfL z-scores were computed as described previously.^25^ Pearson and partial Spearman correlations between baseline NEV synaptopodin z-scores and NfL z-scores were computed to assess unadjusted and adjusted correlations (for age, sex, race, MS subtype, and treatment arm where applicable). The OR and HR models for disability progression in PwMS with both biomarkers were run as described above. To ensure cross-cohort comparability prior to meta-analyses, whole-brain atrophy rates were standardized within each cohort. Linear regression analyses were then conducted following the same approach described above. All models were fitted separately for PMS, PPMS, and SPMS. For the meta-analyses, cohort-specific effect estimates were pooled using fixed-effects inverse-variance-weighted meta-analysis. For dual-biomarker Kaplan-Meier analyses, participants were stratified into four groups based on the combination of NEV synaptopodin and NfL levels, using their respective Youden-derived cutoffs. The cutoff for NEV synaptopodin was as defined above and a cutoff for NfL was computed in the same manner. Overall group differences were assessed using the log-rank test, and a pairwise comparison was performed between the two most discordant groups (NfL↑Synpo↓ vs. NfL↓Synpo↑).

To assess the impact of adding biomarkers on model discrimination, AUC (from logistic regression), Harrell’s C-index (from Cox regression), and partial R² (for brain atrophy) were compared across four model constellations: base model (age, sex, race, baseline EDSS/brain volume, follow-up time and treatment arm in the discovery cohort), base+NEV synaptopodin, base+NfL, and base+both biomarkers. Incremental contributions for nested models were assessed by likelihood-ratio test. These analyses were restricted to SPMS participants with both biomarkers measured.

Confirmed disability progression was the primary endpoint of interest, tested in the discovery cohort and independently replicated in the validation cohort. Benjamini Hochberg correction was applied to pairwise group comparisons. The remaining analyses are exploratory, with nominal p-values reported and no adjustment for multiple comparisons unless stated otherwise. Consistency of effect direction and magnitude across two independent cohorts was used as the principal safeguard against false-positive findings. Unless otherwise stated, tests were two-sided with α = 0.05.

Analyses were performed in R version 4.3.1 (R Core Team) using lme4 for linear mixed-effects models, survival and survminer for Cox regression and Kaplan Meier analyses, pROC for ROC analysis and cutoff derivation, emmeans and car for adjusted group comparisons, and ggplot2 for visualization.

## Data availability

The data supporting the findings of this study are not publicly available because they contain protected health information. De-identified participant data is available from the corresponding authors upon reasonable request, subject to institutional review board approval and data use agreements. Data from the SPRINT-MS trial (NCT01982942) are additionally subject to trial data sharing policies. Source data are provided in the supplementary tables.

## Code availability

No custom code was generated for this study. All analyses were performed in R (v4.3.1) using the publicly available packages as described above.

## Acknowledgments

Funding for the discovery cohort was provided by a DoD IIRA to P.B. Funding for the validation cohort was provided by Genentech to E.M.M. and P.A.C., and by NIH/NINR R01 NR018851 (HEAL) to E.M.M. L.J. was funded by a National MS Society postdoctoral fellowship (NMSS, FG-2511-45413) and a clinician scientist fellowship by the CRC-TRR274 (DFG, ID 408885537). During the preparation of this manuscript, the authors used Claude Opus 4.5 (Anthropic) and Grammarly to improve the language, grammar, and readability of the text. The authors reviewed and edited all output and take full responsibility for the manuscript’s content.

## Author contributions

P.B. conceived the study. P.B. and L.J. designed the study. M.K.S.K., D.J., V.M., R.E., R.T., and L.J. performed NEV isolation, characterization, and synaptopodin quantification. L.J. performed the statistical analyses with contributions from J.G.H., K.C.F., F.S., and M.D.S. L.J. generated the figures. B.E.D., A.G.F., and K.C.F. developed and applied the MRI processing pipeline and provided imaging measures. K.C.F., E.M.M., P.A.C., E.S.S., and S.S. contributed clinical data and participant assessments. P.B., P.A.C., and E.M.M. obtained funding and provided cohort access and resources. C.H. and I.A. contributed to cohort resources, data interpretation, and manuscript revision. L.J. wrote the original draft. P.B. supervised the study. All authors reviewed and edited the manuscript and approved the final version.

