## Supplementary information for "A Blood-Based Synaptic Biomarker in Neuronally-Enriched Extracellular Vesicles Predicts Disability Progression in Progressive Multiple Sclerosis"

**Supplementary Figures and Supplementary Figure Legends**

Supplementary Fig. S1.:

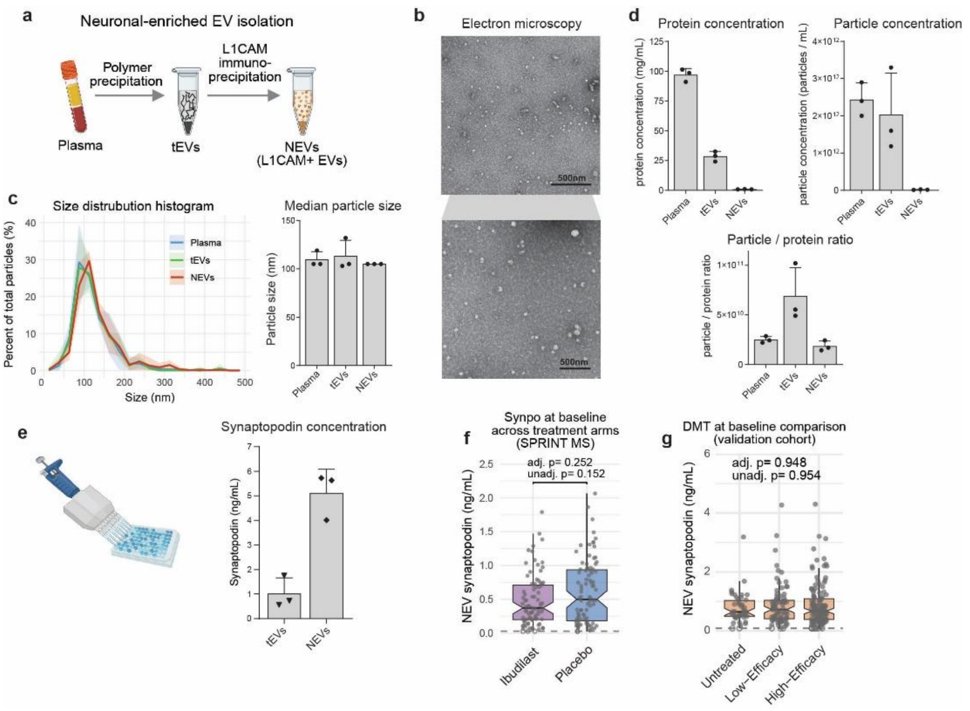


**Supplementary Fig. S1.: NEV isolation workflow, characterization, and baseline NEV synaptopodin comparisons by treatment. (a)** Schematic overview of NEV isolation workflow: including defibrinization of plasma using thrombin, followed by polymer precipitation to enrich plasma particles including extracellular vesicles, and immunoprecipitation using biotinylated L1CAM antibodies and streptavidin beads. **(b)** Transmission electron microscopy (TEM) images of isolated NEVs with negative staining. **(c)** Size distribution histograms of particles in plasma, total EVs (tEVS), and NEVs measured by nanoparticle tracking analysis (NTA). **(d)** Bar plots of the protein concentration (measured with BCA) and particle concentration (measured by NTA) with the particle/protein ratio in plasma, tEVs, and NEVs. **(e)** Bar plot of the synaptopodin concentration in tEVs and NEVs. **(f-g)** Notched boxplots of the baseline NEV synaptopodin concentration by (f) treatment arm in the discovery cohort and (g) DMT treatment in the validation cohort. Unadjusted comparisons used Wilcoxon rank-sum tests for panel f and Kruskal-Wallis tests for panel g. Adjusted p-values are from ANCOVA of synaptopodin z-scores adjusting for age, sex, race, MS/PMS subtype.

Supplementary Fig. S2.:

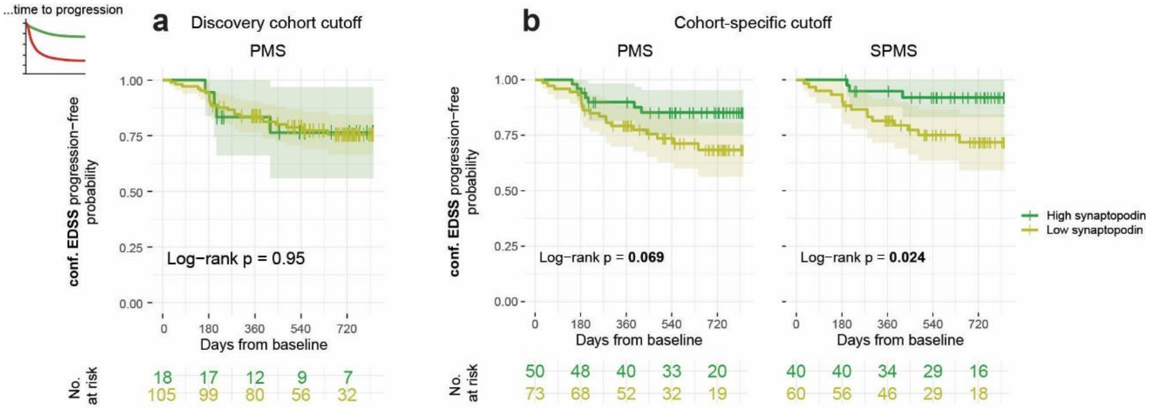


**Supplementary Fig. S2.: Kaplan-Meier curves for the progression-free probability stratified by baseline NEV synaptopodin** (high/dark green vs low/light green) using **(a)** the ROC-derived Youden cutoff from the discovery cohort and **(b)** a ROC-derived Youden cutoff optimized for confirmed EDSS progression and the PMS participants from within the same (validation) cohort. Shaded bands represent 95% confidence intervals; number-at-risk tables and log-rank p-values are shown below.

Supplementary Fig. S3.:

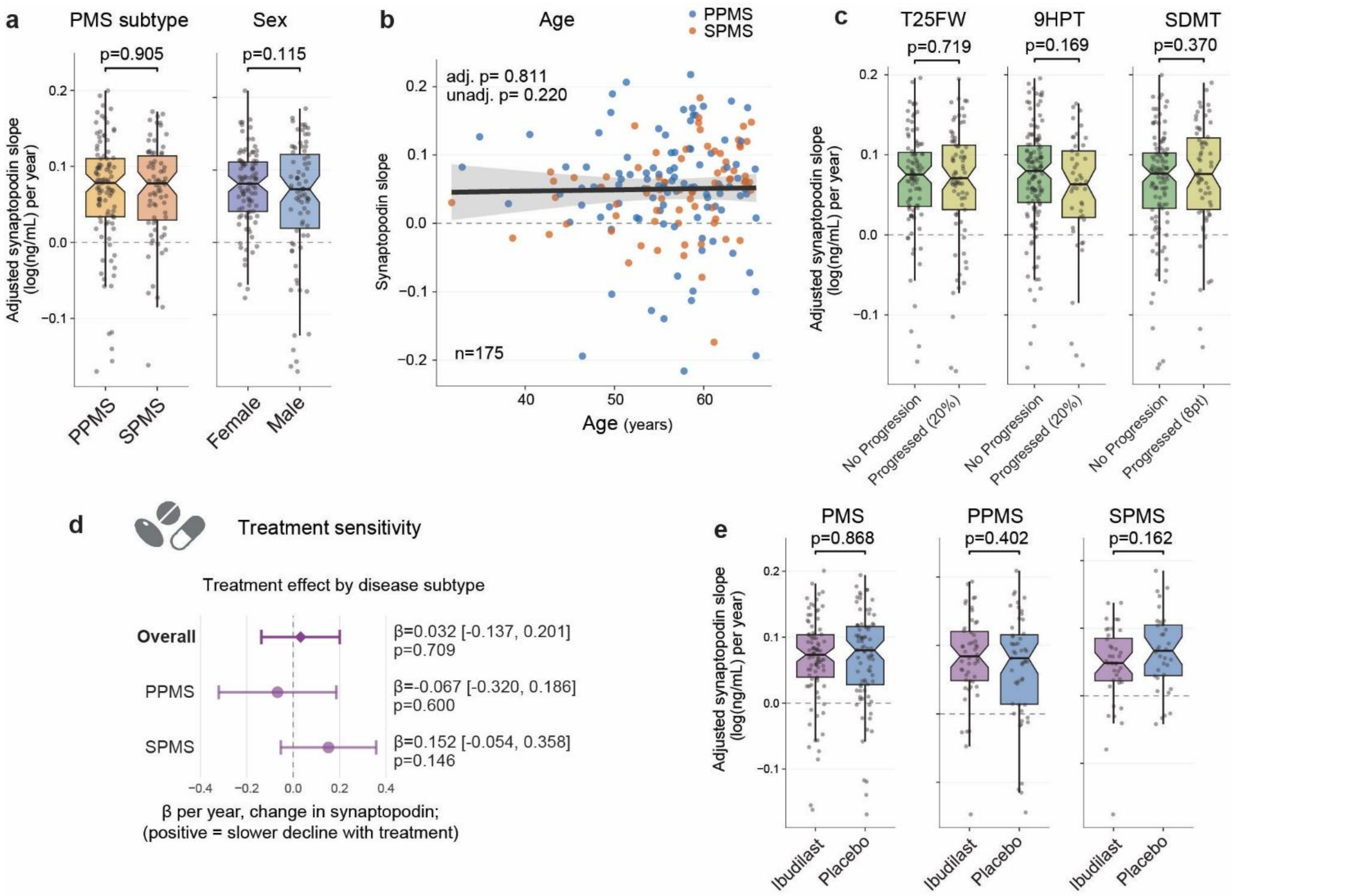


**Supplementary Fig. S3.: Longitudinal NEV synaptopodin change by demographics, association with extended clinical outcomes, and change with ibudilast treatment. (a)** Notched boxplots of adjusted NEV synaptopodin slope [log(ng/mL)/year] by PMS subtype and sex in the discovery cohort. Individual data points are shown as jittered dots. Unadjusted comparisons used Wilcoxon rank-sum tests. Adjusted p-values are from ANCOVA models adjusting for age, sex, race, PMS subtype, treatment arm, and baseline NEV synaptopodin z-score. **(b)** Scatterplot of the association between NEV synaptopodin slope [log(ng/mL)/year] and age in the discovery cohort, with PPMS shown in blue and SPMS in orange. The line and band (95% CI) show the covariate-adjusted association between age and synaptopodin slope, averaged over PMS subtype. The adjusted p is adjusted for the same covariates as in panel a. **(c)** Notched boxplots of adjusted NEV synaptopodin slope by T25FW, 9HPT, and SDMT progression status in PMS participants. Individual data points are shown as jittered dots. Adjusted p-values are adjusted for the same covariates as in panel a, and baseline clinical outcomes. **(d)** Forest plot showing the estimated treatment effect of ibudilast on NEV synaptopodin change over time, derived from linear mixed-effects models run on the full cohort and PPMS/SPMS subgroups (β is the difference in annual NEV synaptopodin change between treatment arms). Covariates included age, sex, race, and MS subtype (for the full cohort model). The x-axis shows the β coefficient, where a positive value indicates a greater annual increase in synaptopodin in the Ibudilast-treated group than in the placebo group. **(e)** Notched boxplots of adjusted NEV synaptopodin slope by treatment arm (Placebo vs. Ibudilast), shown separately for all PMS, PPMS, and SPMS subgroups. Adjusted p-values are from ANCOVA models adjusted for age, sex, race, PMS subtype (as applicable), and baseline NEV synaptopodin.

**Supplementary Tables**

Supplementary Table S1: Baseline NEV synaptopodin across demographics

Supplementary Table S2: Baseline NEV synaptopodin logistic (OR) and cox (HR) models (and extended table: logistic and cox models including participants with 2 missing EDSS functional scores in the validation cohort)

Supplementary Table S3: Association of baseline NEV synaptopodin with clinical worsening slopes (table 3.1) and MRI slopes (table 3.2)

Supplementary Table S4: NEV synaptopodin change over time across demographics

Supplementary Table S5: Association of a change in NEV synaptopodin with clinical worsening slopes

Supplementary Table S6: Association of a change in NEV synaptopodin with disability worsening and MRI slopes

Supplementary Table S7: Ibudilast treatment effect on NEV synaptopodin change over time

Supplementary Table S8: Comparing baseline NfL and NEV synaptopodin as biomarkers predicting clinical progression (8.1) and MRI brain atrophy (8.2)

Supplementary Table S9: Incremental discrimination and model fit of NEV synaptopodin and NfL added to a clinical base model in SPMS
